# The Concurrent Initiation of Medications Is Associated with Discontinuation of Buprenorphine Treatment for Opioid Use Disorder

**DOI:** 10.1101/2020.01.15.20017715

**Authors:** Pengyue Zhang, Chien-Wei Chiang, Sara Quinney, Macarius Donneyong, Bo Lu, Lei Frank Huang, Feixiong Cheng

## Abstract

**Introduction:** Retention in buprenorphine treatment for opioid use disorder (OUD) yields better opioid abstinence and reduces all-cause mortality for patients with OUD. Despite significant efforts have been made to expand the availability and use of buprenorphine in the United States, its retention rates remain on a low level. The current study examines discontinuation of buprenorphine with respect to concurrent initiation of other medications using real-world evidence.

**Methods:** Case-crossover study was conducted to examine discontinuation of buprenorphine using a large-scale longitudinal health dataset including 148,306 commercially-insured individuals initiated on medications for opioid use disorder (MOUD). Odds ratios and Bonferroni adjusted p-values were calculated for medications and therapeutic classes of medications.

**Results:** Clonidine was associated with increased discontinuation risk of buprenorphine both using the buprenorphine dataset alone (OR = 1.583 and adjusted p-value = 1.22 × 10^−6^) and using naltrexone as a comparison drug (OR = 2.706 and adjusted p-value = 4.11 × 10^−5^). Opioid medications (oxycodone, morphine and fentanyl) and methocarbamol were associated with increased discontinuation risk of buprenorphine using the buprenorphine dataset alone (adjusted p-value < 0.05), but not significant using naltrexone as a comparison drug. 6 drug therapeutic classes were associated with increased discontinuation risk of buprenorphine both using the buprenorphine dataset alone and using naltrexone as a comparison drug (adjusted p-value < 0.05).

**Conclusion:** Concurrent initiation of medications is associated with increased discontinuation risk of buprenorphine. Opioid medications are prescribed among patients on MOUD and associated with increased discontinuation risk of buprenorphine. Analgesics is associated with increased discontinuation risk of buprenorphine for patients without previous exposure of pain medications.

## INTRODUCTION

Currently, opioid use disorder (OUD) is recognized as one of the most serious public health concerns in the United States (US). OUD affects as many as 2.1 million Americans and causes up to 43,600 overdose deaths each year (SAMHSA, 2017). The pharmacologic treatment for OUD is through the use of medications for opioid use disorder (MOUD) including methadone, buprenorphine, and naltrexone (SMAHSA, 2019). Evidence-based research demonstrate that retention in buprenorphine yields better opioid abstinence and reduces all-cause mortality for patients with OUD (Mattick, Breen, Kimber, & Davoli, 2014; Sordo et al., 2017). Despite significant efforts have been made to expand the availability and use of buprenorphine in the past decade (Alderks, 2017; Littrell, 2017), its retention rates remain on a low level (Feelemyer, Des Jarlais, Arasteh, Abdul-Quader, & Hagan, 2014). A review of randomized clinical trials (RCTs) identifies a wide range of retention rates (e.g. 6-month retention rates: 3% - 88%) (Thomas et al., 2014). Alternatively, real-world evidence (e.g. large-scale health data) suggest 6-month retention rates are less than 30% to 50% (Morgan, Schackman, Leff, Linas, & Walley, 2018; Samples, Williams, Olfson, & Crystal, 2018; Shcherbakova, Tereso, Spain, & Roose, 2018). As existing data demonstrate that the rates of relapse to illicit opioid exceeded 50% after discontinuation of buprenorphine (Bentzley, Barth, Back, & Book, 2015), a crucial component to address the opioid epidemic is to prevent premature discontinuation of buprenorphine.

Large-scale health data (e.g. US nationwide health insurance claims) are extremely valuable for investigating discontinuation of buprenorphine, as many of the OUD patient groups are under-represented in RCTs (Franklin & Schneeweiss, 2017; Wang et al., 2019). For instance, a literature review of buprenorphine studies shows that only 5 out of 17 RCTs have sample sizes over 200 (Thomas et al., 2014), while OUD affects over 2 million Americans. In large-scale health data, discontinuation of buprenorphine can be determined by patients’ supply of buprenorphine. For instance, discontinuation may be defined as patients being without buprenorphine supply over a period (e.g. 60 days) after initiation (Saloner, Daubresse, & Caleb Alexander, 2017). Subsequently, risk factors for discontinuation of buprenorphine are investigated. These studies include Lopian, Chebolu, Kulak, Kahn, & Blondell (2019), Manhapra, Agbese, Leslie, & Rosenheck (2018), Morgan et al. (2018), Saloner et al. (2017), Samples et al. (2018), and Shcherbakova et al. (2018) etc. Risk factors in these studies include demographic variables, administrative factors (e.g. type of insurance), comorbidities (e.g. depression disorder), and medications. However, the transient effects of concurrent initiation of medications on discontinuation of buprenorphine have not been explicitly investigated. For instance, medications are only measured in the baseline period before initiation of buprenorphine in Samples et al. (2018). Shcherbakova et al. (2018) examines concomitant medications regardless of the concomitant medications’ initiation times. In other words, concomitant medications are treated as binary variables. A variable (i.e. a medication) equals to 1, if the medication has been co-administered with buprenorphine.

Many of OUD patients have physical and/or mental health conditions (e.g. comorbidities) that necessitate treatment with medications (Mark, Dilonardo, Vandivort, & Miller, 2013), while the effects of concurrent initiation of medications on discontinuation of buprenorphine have not been explicitly examined using real-world evidence. In fact, concurrent initiation of medications has high potential to interact with buprenorphine and alter the response of buprenorphine (McCance-Katz, Sullivan, & Nallani, 2010; Moody, Fu, & Fang, 2018; WHO, 2009). For instance, more than 300 drugs listed in Micromedex may interact with buprenorphine (IBM, 2019a). An example is that concomitant use of atazanavir (a HIV treatment) with buprenorphine has been found to significantly alter buprenorphine’s drug response (McCance-Katz et al., 2010). Moreover, when buprenorphine and benzodiazepines (e.g. alprazolam) have been administered together, deaths have resulted that are thought to be related to depression of the central nervous system (CNS) (Schuman-Olivier et al., 2013). Although the aforementioned studies identified important risk factors for discontinuation of buprenorphine, none of them explicitly investigated discontinuation of buprenorphine with respect to concurrent initiation of other medications.

In the current study, we investigate discontinuation of buprenorphine treatment with respect to concurrent initiation of other medications using a large-scale health database and case-crossover design. Additionally, we investigate discontinuation of buprenorphine with respect to the medications’ therapeutic classes [e.g. Anatomical Therapeutic Chemical (ATC) codes (WHO, 2019)]. ATC system is recognized worldwide as a standard to define drug indications, which is related with OUD’s comorbidities. The case-crossover design is developed to study short-term exposures on the risk of acute outcomes (e.g. myocardial infarction) (Maclure, 1991). It is a rigorous, reproducible, controlled epidemiologic study design. Recently, the case-crossover design has been effectively used to identify concurrent initiation of medications that alter the response of a pre-administered drug (Bykov, Mittleman, Glynn, Schneeweiss, & Gagne, 2019; Bykov, Schneeweiss, Glynn, Mittleman, & Gagne, 2019).

## METHODS

### Data preparation

This study utilized the MarketScan commercial claims data (IBM, 2019b) from 2012 to 2017. The dataset included individual-level diagnosis codes, procedure codes and pharmacy claims for ∼50 million patients per year. Patients with at least one diagnosis code related to OUD (Appendix Table 1) and initiated on buprenorphine were included. Specifically, pharmacy prescription of buprenorphine was identified by the definitions in Morgan et al. (2018) and Samples et al. (2018). Clinic-administered buprenorphine was identified by the Healthcare Common Procedure Coding System (HCPCS) codes including J0571, J0572, J0573, J0574 and J0575. Additionally, patients with least one diagnosis code related to OUD (Appendix Table 1) and initiated on naltrexone were selected as a comparison group. Pharmacy prescription of naltrexone was identified by the definitions in Morgan et al., (2018). Clinic-administered naltrexone was identified by the HCPCS code J2315.

**Table 1.**
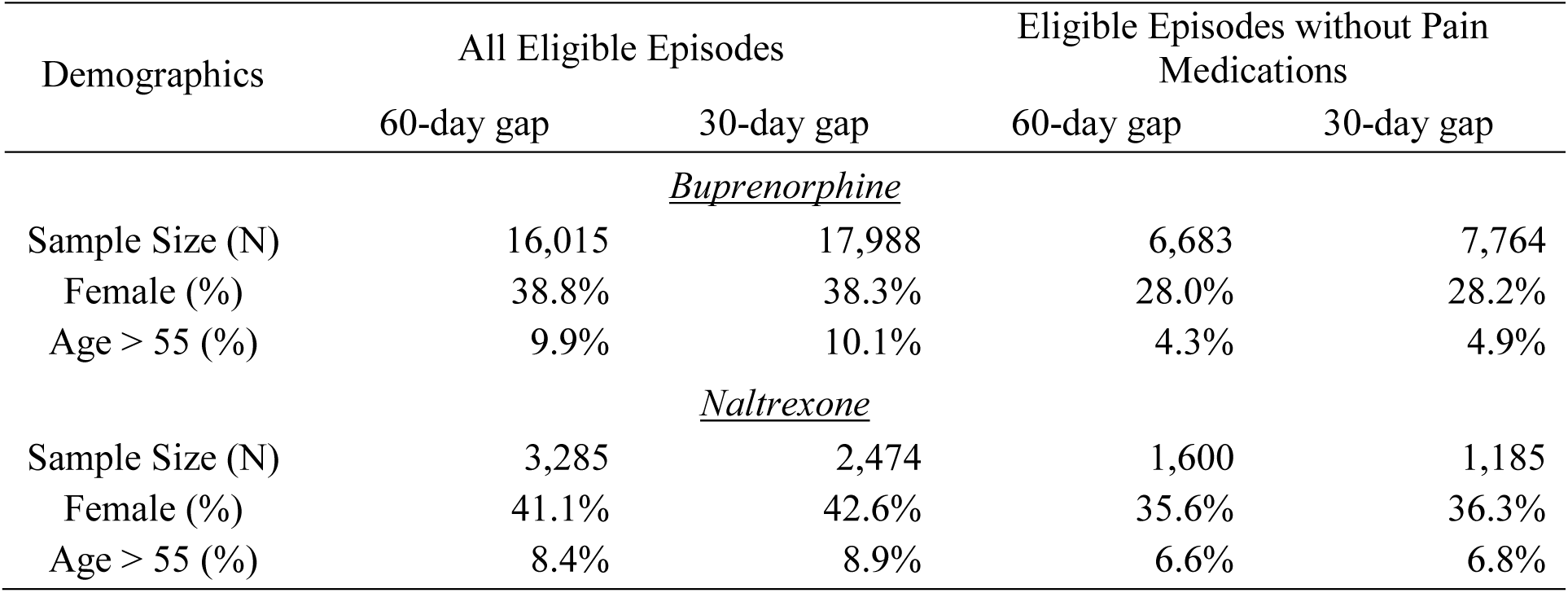
Descriptive statistics for demographic variables

| Demographics | All Eligible Episodes |  | Eligible Episodes without Pain Medications |  |
| --- | --- | --- | --- | --- |
|  | 60-day gap | 30-day gap | 60-day gap | 30-day gap |
| *<u>Buprenorphine</u>* |  |  |  |  |
| Sample Size (N) | 16,015 | 17,988 | 6,683 | 7,764 |
| Female (%) | 38.8% | 38.3% | 28.0% | 28.2% |
| Age > 55 (%) | 9.9% | 10.1% | 4.3% | 4.9% |
| *<u>Naltrexone</u>* |  |  |  |  |
| Sample Size (N) | 3,285 | 2,474 | 1,600 | 1,185 |
| Female (%) | 41.1% | 42.6% | 35.6% | 36.3% |
| Age > 55 (%) | 8.4% | 8.9% | 6.6% | 6.8% |

A MOUD (i.e. buprenorphine and/or naltrexone) episode was defined as the time between initiation and discontinuation. Initiation was defined as the date when MOUD was first supplied (e.g. first prescription). Discontinuation was defined as being without MOUD for a period (e.g. 60 days) based on prescription records or clinic visits. In our analysis, the 60-day gap was used for primary analysis and the 30-day gap was used for sensitivity analysis (Saloner et al., 2017; Samples et al., 2018). We excluded the following episodes: (1) initiated within 180 days of insurance enrollment or a previous episode, (2) duration less than 90 days, (3) with MOUD supply during the last 2 months of insurance enrollment, and (4) received more than 1 type of MOUD. Further, subgroups were selected by excluding baseline exposure to pain medications (e.g. received pain medications before initiation of MOUD). These pain medications (Appendix table 2) were selected according to Guidelines for ATC Classifications and DDD Assignment | 2019 (WHO, 2019). Data preparation are illustrated in figure 1. The demographics for our final datasets are presented in Table 1.

**Table 2.** Drugs and ATC codes associated with discontinuation of buprenorphine using naltrexone as referent.

| ACT level | Drug / ATC code | Exposed in both window | Exposed only in hazardous window | Exposed only in referent window | Odds ratio | Adjusted p-value | Odds ratio using naltrexone as a comparison drug | Adjusted p-value |
| --- | --- | --- | --- | --- | --- | --- | --- | --- |
| *All episodes and 60-day gap for discontinuation* |  |  |  |  |  |  |  |  |
| Drug | clonidine | 383 | 448 | 283 | 1.583 | $1.22 \times 10^{-6}$ | 2.706 | $4.11 \times 10^{-5}$ |
| 2 | antiglaucoma preparations and miotics | 422 | 471 | 305 | 1.544 | $2.97 \times 10^{-6}$ | 2.780 | $1.29 \times 10^{-5}$ |
| 2 | antimigraine preparations | 613 | 565 | 423 | 1.336 | $7.32 \times 10^{-3}$ | 2.388 | $8.85 \times 10^{-5}$ |
| 2 | antihypertensives and diuretics in combination | 428 | 471 | 300 | 1.570 | $8.61 \times 10^{-7}$ | 2.486 | $2.85 \times 10^{-5}$ |
| 2 | antiadrenergic agents, centrally acting | 394 | 452 | 295 | 1.532 | $1.08 \times 10^{-5}$ | 2.747 | $1.03 \times 10^{-5}$ |
| 2 | anxiolytics | 2333 | 874 | 698 | 1.252 | $1.06 \times 10^{-2}$ | 1.704 | $1.12 \times 10^{-3}$ |
| 3 | antihypertensives | 460 | 474 | 313 | 1.514 | $1.12 \times 10^{-5}$ | 2.431 | $2.70 \times 10^{-5}$ |
| *All episodes and 30-day gap for discontinuation* |  |  |  |  |  |  |  |  |
| Drug | clonidine | 389 | 463 | 296 | 1.564 | $1.61 \times 10^{-6}$ | 4.087 | $1.56 \times 10^{-7}$ |
| 2 | antiglaucoma preparations and miotics | 436 | 487 | 328 | 1.485 | $3.05 \times 10^{-5}$ | 4.108 | $1.55 \times 10^{-7}$ |
| 2 | antimigraine preparations | 651 | 597 | 440 | 1.357 | $1.23 \times 10^{-3}$ | 3.155 | $1.24 \times 10^{-6}$ |
| 2 | antihypertensives and diuretics in combination | 444 | 486 | 318 | 1.528 | $3.74 \times 10^{-6}$ | 3.578 | $2.42 \times 10^{-8}$ |
| 2 | antiadrenergic agents, centrally acting | 398 | 471 | 311 | 1.514 | $1.26 \times 10^{-5}$ | 4.212 | $2.34 \times 10^{-8}$ |
| 3 | antihypertensives | 476 | 494 | 337 | 1.466 | $6.15 \times 10^{-5}$ | 3.505 | $1.65 \times 10^{-8}$ |
| *Episodes without pain medications and 60-day gap for discontinuation* |  |  |  |  |  |  |  |  |
| 2 | antiadrenergic agents, centrally acting | 94 | 164 | 97 | 1.691 | $2.78 \times 10^{-2}$ | 3.381 | $4.30 \times 10^{-2}$ |
| 3 | analgesics | 174 | 381 | 226 | 1.686 | $2.60 \times 10^{-7}$ | 2.299 | $4.22 \times 10^{-2}$ |
| *Episodes without pain medications and 30-day gap for discontinuation* |  |  |  |  |  |  |  |  |
| Drug | clonidine | 89 | 183 | 98 | 1.867 | $3.33 \times 10^{-4}$ | 6.769 | $1.73 \times 10^{-3}$ |
| 2 | antiglaucoma preparations and miotics | 95 | 185 | 106 | 1.745 | $3.06 \times 10^{-3}$ | 6.545 | $2.24 \times 10^{-3}$ |
| 2 | antimigraine preparations | 104 | 207 | 118 | 1.754 | $6.68 \times 10^{-4}$ | 5.847 | $3.08 \times 10^{-3}$ |
| 2 | antihypertensives and<br>diuretics in combination | 104 | 187 | 103 | 1.816 | $6.82 \times 10^{-4}$ | 4.798 | $1.14 \times 10^{-3}$ |
| 2 | antiadrenergic agents,<br>centrally acting | 93 | 185 | 104 | 1.779 | $1.59 \times 10^{-3}$ | 7.338 | $4.84 \times 10^{-4}$ |
| 2 | analgesics | 179 | 436 | 249 | 1.751 | $7.57 \times 10^{-10}$ | 2.711 | $1.51 \times 10^{-2}$ |
| 3 | antihypertensives | 112 | 189 | 111 | 1.703 | $5.63 \times 10^{-3}$ | 4.986 | $4.54 \times 10^{-4}$ |

**Figure 1.**
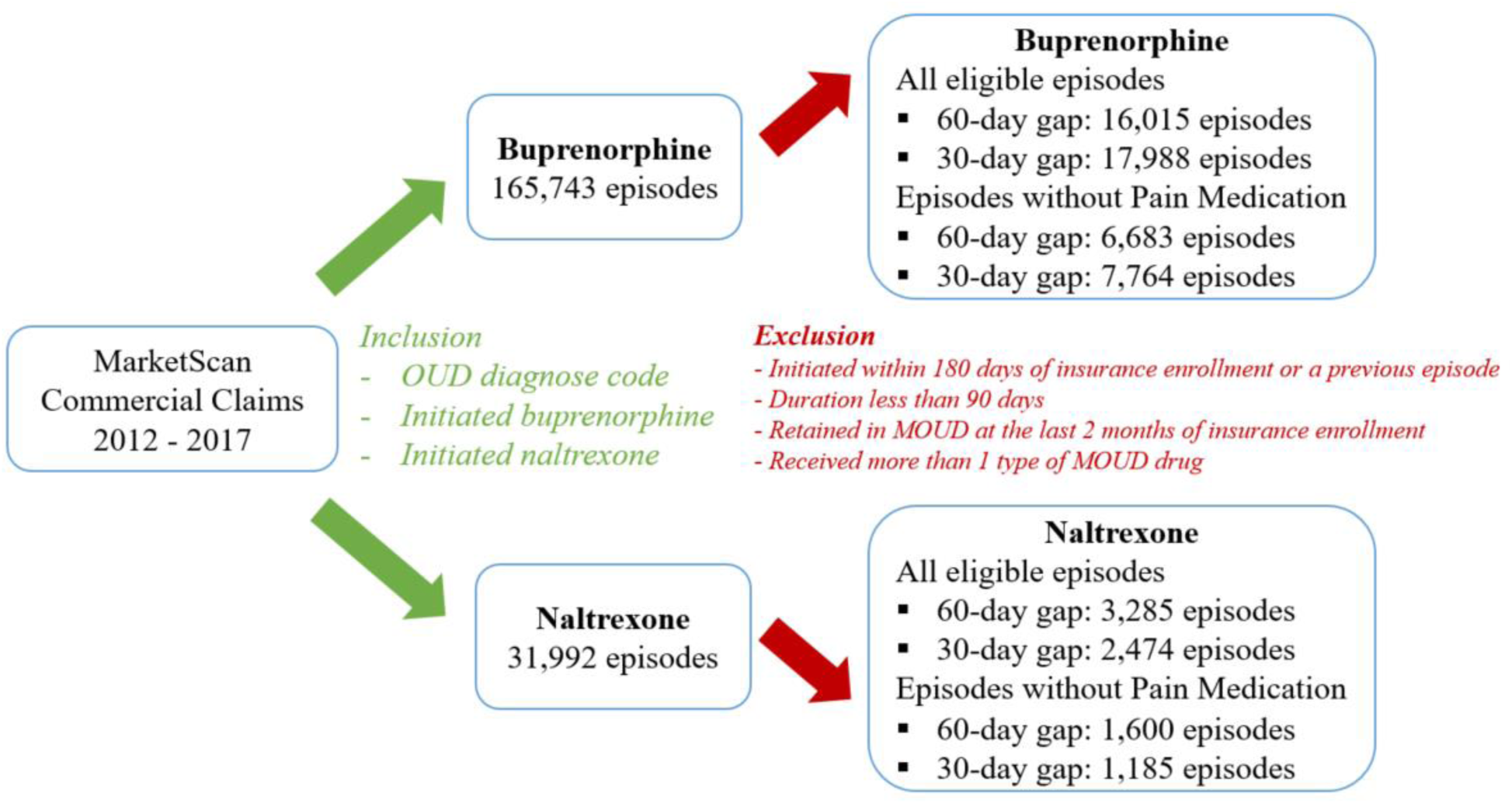
Overview of data preparation.

### Case-crossover design and statistical Analysis

The case-crossover design is depicted in figure 2. The referent window, washout period, and hazardous window were determined to be 30-day, which was suggested for investigating the effects of concurrent initiation of medications (Bykov, Mittleman, et al., 2019; Bykov, Schneeweiss, et al., 2019). Exposure to concomitant medications was examined in the referent window (61 – 90 days before discontinuation) and hazardous window (1 – 30 days before discontinuation). Further, the concomitant medications were mapped to their generic names using RxNorm (NLM, 2019), compounded medications were mapped to generic names of their components, and all medications were mapped to their corresponding ATC codes (level 2 – 3).

**Figure 2.**
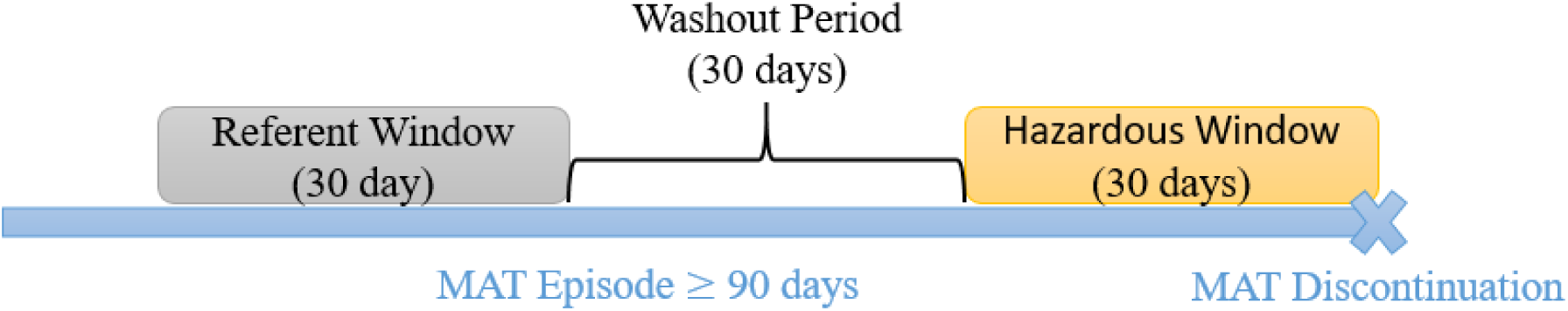
Case-crossover design.

We conducted case-crossover design for all datasets (figure 1). For a medication or an ATC code, the outcomes were: a) the number of episodes exposed to the medication (or the ATC code) in the referent window, but unexposed in the hazardous window, and b) the number of episodes unexposed in the referent window, but exposed in the hazardous window. We calculated the odds ratio and Bonferroni adjusted p-value (McNemar test) for each medication and ATC code using the buprenorphine dataset alone. Additionally, we calculated the odds ratio for buprenorphine discontinuation using naltrexone as a comparison drug, as well as the Bonferroni adjusted p-value (normal approximation). In another word, by using naltrexone as a comparison drug, the analysis utilizes both the buprenorphine dataset and the naltrexone dataset.

## Results

In this analysis, we computed the odds ratio and p-value for each of the medications, ATC level 2 codes, and ATC level 3 codes. Additionally, we computed the odds ratios and p-values for buprenorphine discontinuation using naltrexone as a comparison drug. Further, for each of the 4 buprenorphine datasets, p-values were adjusted by the total number of tests (e.g. the total number of medications and ATC codes). The drugs and ATC codes with adjusted p-values less than 0.05 using naltrexone as a comparison drug are presented in table 2. For all episodes, the results are consistent between two definitions of discontinuation (i.e. 60-day gap and 30-day gap). We identified 7 associations from the 60-day gap analysis. Specifically, clonidine was associated with increased discontinuation risk of buprenorphine. For the buprenorphine dataset alone, clonidine had odds ratio equal to 1.583 (adjusted p-value = 1.22 × 10^−6^). Using naltrexone as a comparison drug, the odds ratio increased to 2.706 (adjusted p-value = 4.11 × 10^−5^). Five ATC level 2 codes and the ATC level 3 code antihypertensives were associated with increased discontinuation risk of buprenorphine (adjusted p-value < 0.05). Using episodes without pain medications and 60-day gap as the definition of discontinuation, the ATC level 2 code antiadrenergic agents (centrally acting) and the ATC level 3 code analgesics were associated with increased discontinuation risk of buprenorphine (adjusted p-value < 0.05). Using episodes without pain medications and 30-day gap as the definition of discontinuation, clonidine and six ATC codes were associated with increased discontinuation risk of buprenorphine (adjusted p-value < 0.05).

For the buprenorphine dataset alone, using all episodes and 60-day gap as the definition of discontinuation, 4 medications were associated with increased discontinuation risk of buprenorphine. Specifically, oxycodone had odds ratio equal to 1.681 (adjusted p-value = 8.07 × 10^−10^) using the buprenorphine dataset alone, while the odds ratio equaled to 1.107 (adjusted p-value = 1) using naltrexone as a comparison drug. Morphine (odds ratio = 3.613 and p-value = 1.47 × 10^−10^) and fentanyl (odds ratio = 6.400 and p-value < 1 × 10^−10^) exposure were only observed in the buprenorphine dataset. Methocarbamol had odds ratio equal to 1.737 (adjusted p-value = 2.31 × 10^−3^) using the buprenorphine dataset alone, while the odds ratio equaled to 2.217 (adjusted p-value = 1) using naltrexone as a comparison drug. The ACT level 3 code opioids had odds ratio equal to 1.663 (adjusted p-value < 1 × 10^−10^) using the buprenorphine dataset alone, while the odds ratio equaled to 0.998 (adjusted p-value = 1) using naltrexone as a referent drug. Similar results for methocarbamol and the ACT level 3 code opioids were observed using episodes without pain medications and 60-day gap as the definition for discontinuation.

## Discussion

This study investigates discontinuation of buprenorphine with respect to concurrent initiation of other medications using a US nationwide insurance claim dataset. Specifically, we investigate medications and the drug therapeutic classes (e.g. ATC code) for discontinuation of buprenorphine. We identify clonidine and six ATC codes that associated with increased discontinuation risk of buprenorphine both using the buprenorphine dataset alone and using naltrexone as a comparison drug. All of these associations have Bonferroni adjusted p-values less than 0.05. Additionally, we identify medications that are associated with increased discontinuation risk of buprenorphine using the buprenorphine dataset alone, but not statistically significant using naltrexone as a comparison drug. To our knowledge, this is the first large-scale study that investigate discontinuation of buprenorphine with respect to the management of OUD’s comorbidity by initiating other medications. Our findings can be used to reduce premature discontinuation of buprenorphine.

First, we would like to point out that opioid medications are associated with increased discontinuation risk of buprenorphine, although not statistically significant using naltrexone as a comparison drug (table 3). In our analysis, many of the patients on MOUD are prescribed with codeine, fentanyl, hydrocodone, morphine, oxycodone and tramadol; and similar statistics are observed by Shcherbakova et al. (2018). Overprescribing of opioid medications is believed to be a major cause for the current opioid epidemic (Manchikanti et al., 2012). For patients on MOUD, a switch from buprenorphine to other opioids may be planned at the time of a surgical procedure to manage post-operative pain. Although the relationships between prescription opioid pain medications and discontinuation of buprenorphine have not been rigorously investigated, it should be recognized that opioid pain medications are associated with increased discontinuation risk of buprenorphine and are prescribed among patients on MOUD. Given the existence of prescription drug monitoring programs (PDMPs) in virtually all US states (Haffajee, Jena, & Weiner, 2015; Strickler et al., 2019), there are opportunities to track opioid prescriptions for patients on MOUD and to make appropriate changes in treatment plans for reducing premature MOUD discontinuation.

**Table 3.** Drugs and the ATC code opioid associated with discontinuation of buprenorphine.

| ACT level | Drug / ATC code | Exposed in both window | Exposed only in hazardous window | Exposed only in referent window | Odds ratio | Adjusted p-value | Odds ratio using naltrexone as referent | Adjusted p-value |
| --- | --- | --- | --- | --- | --- | --- | --- | --- |
| *All episodes and 60-day gap for discontinuation* |  |  |  |  |  |  |  |  |
| Drug | oxycodone | 694 | 501 | 298 | 1.681 | $8.07 \times 10^{-10}$ | 1.107 | 1 |
| Drug | morphine | 40 | 112 | 31 | 3.613 | $1.47 \times 10^{-10}$ | NA | NA |
| Drug | fentanyl | 23 | 128 | 20 | 6.400 | $< 1 \times 10^{-10}$ | NA | NA |
| Drug | methocarbamol | 150 | 198 | 114 | 1.737 | $2.31 \times 10^{-3}$ | 2.271 | 1 |
| ACT 3 | opioids | 1234 | 853 | 513 | 1.663 | $< 1 \times 10^{-10}$ | 0.998 | 1 |
| *Episodes without pain medications and 60-day gap for discontinuation* |  |  |  |  |  |  |  |  |
| Drug | methocarbamol | 8 | 56 | 17 | 3.294 | $4.21 \times 10^{-3}$ | 2.471 | 1 |
| ACT 3 | opioids | 39 | 136 | 68 | 2.000 | $1.59 \times 10^{-3}$ | 1.556 | 1 |

Second, clonidine is associated with increased discontinuation risk of buprenorphine. Clonidine a centrally acting α2-selective adrenergic receptor agonist used to treat hypertension. According to the guideline of American Society of Addiction Medicine (ASAM), clonidine is recommended to support opioid withdrawal, although it is not US FDA approved for the treatment of opioid withdrawal (ASAM, 2019). For buprenorphine as MOUD, effect of combined use of clonidine and buprenorphine is investigated in a clinical trial (Kowalczyk et al., 2015), in which no group difference in time to opioid relapse is observed between buprenorphine alone and buprenorphine with clonidine (P = 0.05). The discontinuation of buprenorphine associated with initiation of clonidine may due to either pharmacological reasons or clinical reasons. On the pharmacology side, clonidine has the potential to increase opioid abuse by boosting and extending the opioid-related “high” (Seale, Dittmer, Sigman, Clemons, & Johnson, 2014). Additionally, combined use of clonidine and buprenorphine may increases risk of severe sedation and other adverse drug events (FDA, 2016; WHO, 2009). As clonidine is associated with increased discontinuation risk of buprenorphine both using buprenorphine dataset alone and using naltrexone as a comparison drug, the possibility of drug-drug interaction between clonidine and buprenorphine deserves further investigation. On the clinical side, discontinuation of buprenorphine with initiation of clonidine may actually be requested by patients to stop or reduce the dosage of buprenorphine treatment. Careful management is required for their co-administration.

Third, methocarbamol is associated with increased discontinuation risk of buprenorphine using the buprenorphine dataset alone. Although the odds ratio increases to 2.217 using naltrexone as a comparison drug, the adjusted p-value = 1 due to smaller sample size of the naltrexone dataset. Methocarbamol is a CNS depressant with sedative and musculoskeletal relaxant properties. The discontinuation of buprenorphine associated with initiation of methocarbamol may be caused by pharmacological reasons. Methocarbamol has the potential to alter buprenorphine’s buprenorphine’s metabolism by inhibiting CYP3A4, which is buprenorphine’s metabolizer (Moody et al., 2018). Additionally, combined use of methocarbamol and buprenorphine may yield severe adverse reaction due to depression of the CNS (FDA, 2016). Thus, careful management is required for their co-administration.

Forth, using all episodes and 60-day gap as the definition for discontinuation, six ATC codes are associated with increased discontinuation risk of buprenorphine. Five out of the six ATC codes include clonidine. They are antiglaucoma preparations and miotics, antimigraine preparations, antihypertensives and diuretics in combination, antiadrenergic agents centrally acting, and antihypertensives. Thus, these associations may be dominated by clonidine. The ATC level 2 code anxiolytics doesn’t include clonidine. In fact, many of the drugs belonging to anxiolytics are benzodiazepines (WHO, 2019). Thus, the discontinuation of buprenorphine associated with anxiolytics may due to severe depression of CNS (FDA, 2016; WHO, 2009). For episodes without pain medications, analgesics is strongly associated with buprenorphine discontinuation using buprenorphine dataset alone (p-value = 2.60 × 10^−7^), and barely significant using naltrexone as a comparison drug (p-value = 4.22 × 10^−2^). As analgesics is not significant in the all episodes analysis, “new” exposure of analgesics may increase the risk of buprenorphine discontinuation. Our finding suggests that treating OUD’s comorbidities may alter buprenorphine’s response. In healthcare practice, physicians who prescribe MOUD are likely to be different from those that engage in chronic care management. For instance, OUD patients often attend addiction treatment programs which are separate from their primary care physicians or other mental health providers. Indeed, the vast majority of clinicians are not waivered to prescribe buprenorphine (Hutchinson, Catlin, Andrilla, Baldwin, & Rosenblatt, 2014). Such a fragmentation in the system increases the likelihood of premature discontinuation of buprenorphine due to concurrent initiation of other medications.

This study has some limitations. Although our dataset contains a geographically diverse population of commercially-insured Americans, the results are not representative of individuals who are not commercially-insured or uninsured. Additionally, individuals initiated MOUD without an OUD diagnosis are excluded from our analysis to improve the sensitivity of MOUD. There are some limitations with respect to the case-crossover design. First, it requires the patients to be continuously on MOUD over a period (e.g. 90 days). Consequently, the results are not representative of individuals who discontinued in less than 90 days. Last, case-crossover design is subject to time varying confounding.

To summarize, we investigate the discontinuation risk of buprenorphine with respect to medications and ATC codes. This study generates important knowledge for managing OUD patients’ comorbidities, while they are receiving buprenorphine as MOUD. Our results offer the great promise to improve retention and prevent necessary discontinuation of buprenorphine. Last, we would like to point out two important future research directions including extent the current analysis to a broader population (e.g. the Medicaid population) and upscale the current analysis with respect to investigate the associations between combinations of medications and discontinuation of MOUD.

## Data Availability

NA

## Acknowledgements

L.F.H is supported by a grant from CancerFree KIDS foundation 2019 and the start-up funding from the Brain Tumor Center, Division of Experimental Hematology and Cancer Biology, Cancer and Blood Disease Institute, Cincinnati Children’s Hospital Medical Center.

---

**Appendix table 1.**
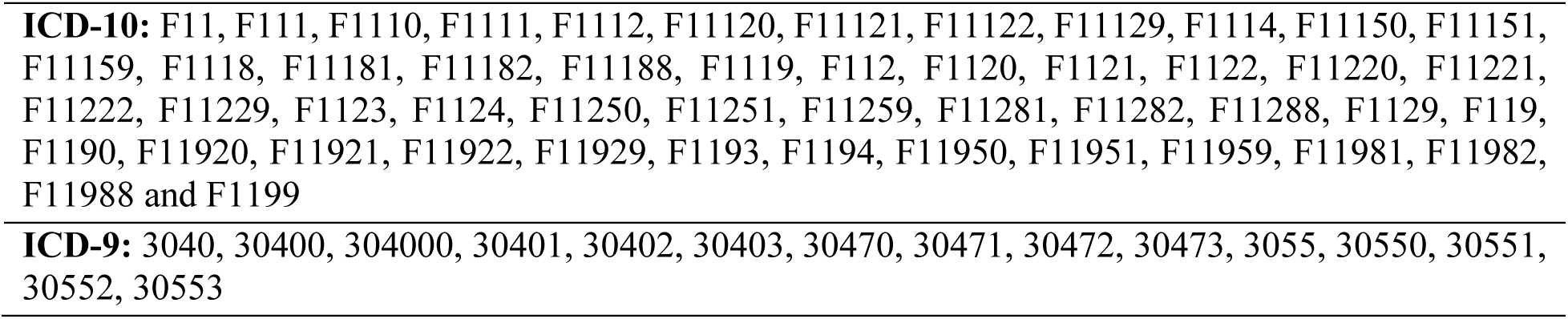
ICD codes for opioid use disorder (OUD)

**Appendix table 2.**
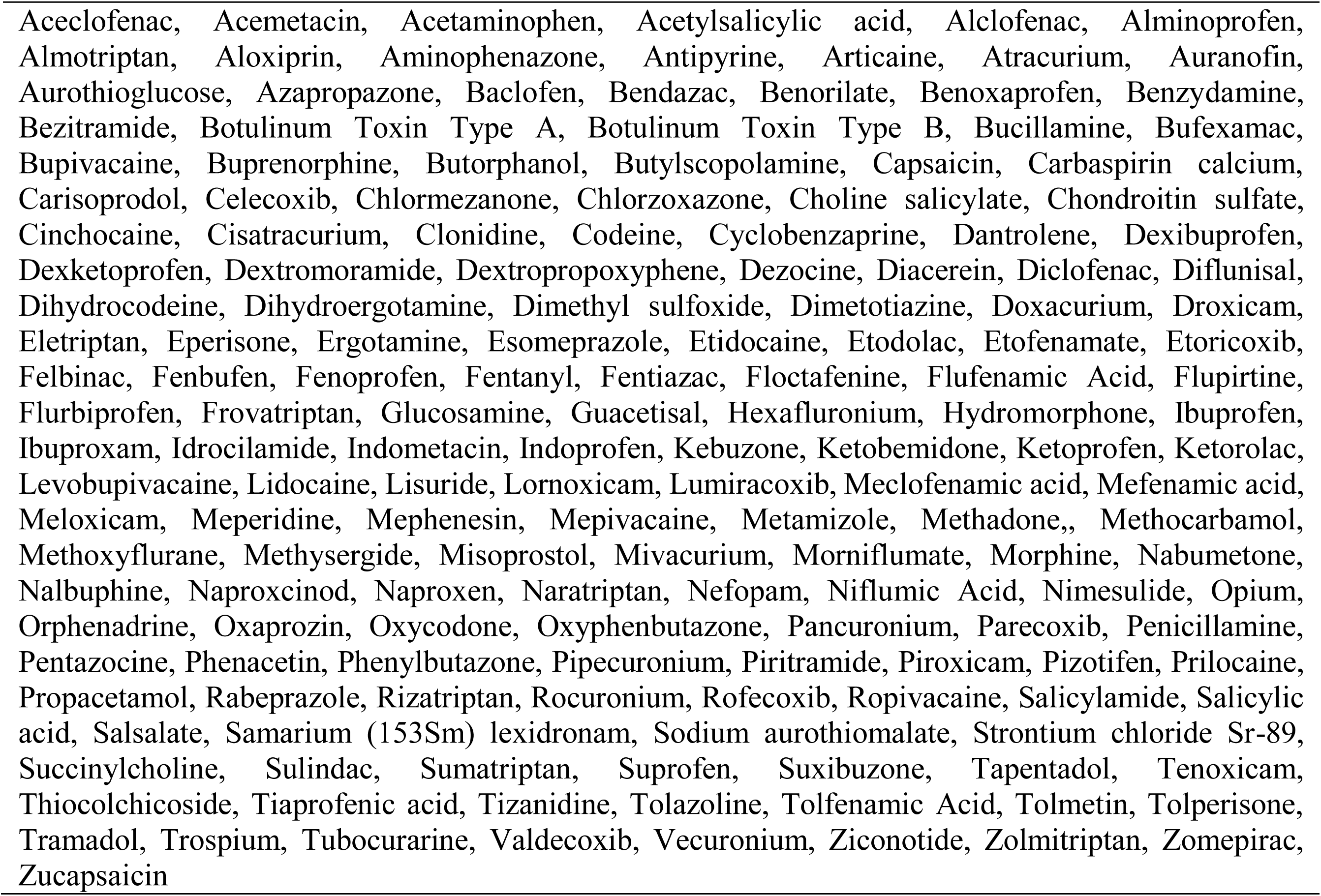
List of pain medications

## REFERENCES

Alderks, C. E. (2017). Trends in the Use of Methadone, Buprenorphine, and Extended-release Naltrexone at Substance Abuse Treatment Facilities: 2003-2015 (Update). Retrieved from https://www.samhsa.gov/data/sites/default/files/report_3192/ShortReport-3192.html

ASAM. (2019). National Practice Guideline for the Use of Medications in the Treatment of Addiction Involving Opioid Use. Retrieved from https://www.asam.org/docs/default-source/practice-support/guidelines-and-consensus-docs/asam-national-practice-guideline-supplement.pdf.

Bentzley, B. S., Barth, K. S., Back, S. E., & Book, S. W. (2015). Discontinuation of buprenorphine maintenance therapy: perspectives and outcomes. J Subst Abuse Treat, 52, 48–57. Retrieved from https://www.ncbi.nlm.nih.gov/pubmed/25601365. doi:10.1016/j.jsat.2014.12.011

Bykov, K., Mittleman, M. A., Glynn, R. J., Schneeweiss, S., & Gagne, J. J. (2019). The Case-Crossover Design for Drug-Drug Interactions: Considerations for Implementation. Epidemiology, 30(2), 204–211. Retrieved from https://www.ncbi.nlm.nih.gov/pubmed/30433922. doi:10.1097/EDE.0000000000000944

Bykov, K., Schneeweiss, S., Glynn, R. J., Mittleman, M. A., & Gagne, J. J. (2019). A Case-Crossover-Based Screening Approach to Identifying Clinically Relevant Drug-Drug Interactions in Electronic Healthcare Data. Clinical Pharmacology & Therapeutics. Retrieved from https://www.ncbi.nlm.nih.gov/pubmed/30663781. doi:10.1002/cpt.1376

FDA. (2016). Drug Safety Communication. Retrieved from https://www.fda.gov/media/107888/download

Feelemyer, J., Des Jarlais, D., Arasteh, K., Abdul-Quader, A. S., & Hagan, H. (2014). Retention of participants in medication-assisted programs in low- and middle-income countries: an international systematic review. Addiction, 109(1), 20–32. Retrieved from https://www.ncbi.nlm.nih.gov/pubmed/23859638. doi:10.1111/add.12303

Franklin, J. M., & Schneeweiss, S. (2017). When and How Can Real World Data Analyses Substitute for Randomized Controlled Trials? Clinical Pharmacology & Therapeutics, 102(6), 924–933. Retrieved from <Go to ISI>://WOS:000414921800020. doi:10.1002/cpt.857

Haffajee, R. L., Jena, A. B., & Weiner, S. G. (2015). Mandatory Use of Prescription Drug Monitoring Programs. Jama-Journal of the American Medical Association, 313(9), 891–892. Retrieved from <Go to ISI>://WOS:000350231800007. doi:10.1001/jama.2014.18514

Hutchinson, E., Catlin, M., Andrilla, C. H. A., Baldwin, L. M., & Rosenblatt, R. A. (2014). Barriers to Primary Care Physicians Prescribing Buprenorphine. Annals of Family Medicine, 12(2), 128–133. Retrieved from <Go to ISI>://WOS:000336799900006. doi:10.1370/afm.1595

IBM. (2019a). Micromedex. Retrieved from https://www.micromedexsolutions.com/micromedex2/librarian/

IBM. (2019b). MarketScan Database. Retrieved from https://marketscan.truvenhealth.com/marketscanportal/

Kowalczyk, W. J., Phillips, K. A., Jobes, M. L., Kennedy, A. P., Ghitza, U. E., Agage, D. A., … Preston, K. L. (2015). Clonidine Maintenance Prolongs Opioid Abstinence and Decouples Stress From Craving in Daily Life: A Randomized Controlled Trial With Ecological Momentary Assessment. Am J Psychiatry, 172(8), 760–767. Retrieved from https://www.ncbi.nlm.nih.gov/pubmed/25783757. doi:10.1176/appi.ajp.2014.14081014

Littrell, J. (2017). Expanding access to Medication Assisted Treatment: The U.S. government’s response to the current heroin epidemic. Social Work in Mental Health, 15(2), 209–229. Retrieved from <Go to ISI>://WOS:000411484000006. doi:10.1080/15332985.2016.1210555

Lopian, K. M., Chebolu, E., Kulak, J. A., Kahn, L. S., & Blondell, R. D. (2019). A retrospective analysis of treatment and retention outcomes of pregnant and/or parenting women with opioid use disorder. J Subst Abuse Treat, 97, 1–6. Retrieved from https://www.ncbi.nlm.nih.gov/pubmed/30577894. doi:10.1016/j.jsat.2018.11.002

Maclure, M. (1991). The case-crossover design: a method for studying transient effects on the risk of acute events. Am J Epidemiol, 133(2), 144–153. Retrieved from https://www.ncbi.nlm.nih.gov/pubmed/1985444. doi:10.1093/oxfordjournals.aje.a115853

Manchikanti, L., Helm, S., 2nd, Fellows, B., Janata, J. W., Pampati, V., Grider, J. S., & Boswell, M. V. (2012). Opioid epidemic in the United States. Pain Physician, 15(3 Suppl), ES9–38. Retrieved from https://www.ncbi.nlm.nih.gov/pubmed/22786464.

Manhapra, A., Agbese, E., Leslie, D. L., & Rosenheck, R. A. (2018). Three-Year Retention in Buprenorphine Treatment for Opioid Use Disorder Among Privately Insured Adults. Psychiatr Serv, 69(7), 768–776. Retrieved from https://www.ncbi.nlm.nih.gov/pubmed/29656707. doi:10.1176/appi.ps.201700363

Mark, T. L., Dilonardo, J., Vandivort, R., & Miller, K. (2013). Psychiatric and medical comorbidities, associated pain, and health care utilization of patients prescribed buprenorphine. Journal of Substance Abuse Treatment, 44(5), 481–487. Retrieved from <Go to ISI>://WOS:000316835700003. doi:10.1016/j.jsat.2012.11.004

Mattick, R. P., Breen, C., Kimber, J., & Davoli, M. (2014). Buprenorphine maintenance versus placebo or methadone maintenance for opioid dependence. Cochrane Database of Systematic Reviews(2). Retrieved from <Go to ISI>://WOS:000332082400003. doi:ARTN CD002207 10.1002/14651858.CD002207.pub4

McCance-Katz, E. F., Sullivan, L. E., & Nallani, S. (2010). Drug Interactions of Clinical Importance among the Opioids, Methadone and Buprenorphine, and Other Frequently Prescribed Medications: A Review. American Journal on Addictions, 19(1), 4–16. Retrieved from <Go to ISI>://WOS:000272864300003. doi:10.1111/j.1521-0391.2009.00005.x

Moody, D. E., Fu, Y. Q., & Fang, W. F. B. (2018). Inhibition of In Vitro Metabolism of Opioids by Skeletal Muscle Relaxants. Basic & Clinical Pharmacology & Toxicology, 123(3), 327–334. Retrieved from <Go to ISI>://WOS:000441237300016. doi:10.1111/bcpt.12999

Morgan, J. R., Schackman, B. R., Leff, J. A., Linas, B. P., & Walley, A. Y. (2018). Injectable naltrexone, oral naltrexone, and buprenorphine utilization and discontinuation among individuals treated for opioid use disorder in a United States commercially insured population. J Subst Abuse Treat, 85, 90–96. Retrieved from https://www.ncbi.nlm.nih.gov/pubmed/28733097. doi:10.1016/j.jsat.2017.07.001

NLM (2019), RXnorm. Retrieved from https://www.nlm.nih.gov/research/umls/rxnorm/

Saloner, B., Daubresse, M., & Caleb Alexander, G. (2017). Patterns of Buprenorphine-Naloxone Treatment for Opioid Use Disorder in a Multistate Population. Med Care, 55(7), 669–676. Retrieved from https://www.ncbi.nlm.nih.gov/pubmed/28410339. doi:10.1097/MLR.0000000000000727

SAMHSA. (2017). 2017 National Survey on Drug Use and Health. Retrieved from https://www.samhsa.gov/data/report/2017-nsduh-annual-national-report

Samples, H., Williams, A. R., Olfson, M., & Crystal, S. (2018). Risk factors for discontinuation of buprenorphine treatment for opioid use disorders in a multi-state sample of Medicaid enrollees. J Subst Abuse Treat, 95, 9–17. Retrieved from https://www.ncbi.nlm.nih.gov/pubmed/30352671. doi:10.1016/j.jsat.2018.09.001

Schuman-Olivier, Z., Hoeppner, B. B., Weiss, R. D., Borodovsky, J., Shaffer, H. J., & Albanese, M. J. (2013). Benzodiazepine use during buprenorphine treatment for opioid dependence: Clinical and safety outcomes. Drug and Alcohol Dependence, 132(3), 580–586. Retrieved from <Go to ISI>://WOS:000325510700026. doi:10.1016/j.drugalcdep.2013.04.006

Seale, J. P., Dittmer, T., Sigman, E. J., Clemons, H., & Johnson, J. A. (2014). Combined Abuse of Clonidine and Amitriptyline in a Patient on Buprenorphine Maintenance Treatment. Journal of Addiction Medicine, 8(6), 476–478. Retrieved from <Go to ISI>://WOS:000345117200014. doi:10.1097/Adm.0000000000000081

Shcherbakova, N., Tereso, G., Spain, J., & Roose, R. J. (2018). Treatment Persistence Among Insured Patients Newly Starting Buprenorphine/Naloxone for Opioid Use Disorder. Ann Pharmacother, 52(5), 405–414. Retrieved from https://www.ncbi.nlm.nih.gov/pubmed/29302989. doi:10.1177/1060028017751913

SMAHSA. (2019). Opioid Dependency Medications. Retrieved from https://www.samhsa.gov/medication-assisted-treatment/treatment#medications-used-in-mat

Sordo, L., Barrio, G., Bravo, M. J., Indave, B. I., Degenhardt, L., Wiessing, L., … Pastor-Barriuso, R. (2017). Mortality risk during and after opioid substitution treatment: systematic review and meta-analysis of cohort studies. Bmj-British Medical Journal, 357. Retrieved from <Go to ISI>://WOS:000400438000001. doi:ARTN j1550 10.1136/bmj.j1550

Strickler, G. K., Zhang, K., Halpin, J. F., Bohnert, A. S. B., Baldwin, G. T., & Kreiner, P. W. (2019). Effects of mandatory prescription drug monitoring program (PDMP) use laws on prescriber registration and use and on risky prescribing. Drug and Alcohol Dependence, 199, 1–9. Retrieved from <Go to ISI>://WOS:000470946100001. doi:10.1016/j.drugalcdep.2019.02.010

Thomas, C. P., Fullerton, C. A., Kim, M., Montejano, L., Lyman, D. R., Dougherty, R. H., … Delphin-Rittmon, M. E. (2014). Medication-assisted treatment with buprenorphine: assessing the evidence. Psychiatr Serv, 65(2), 158–170. Retrieved from https://www.ncbi.nlm.nih.gov/pubmed/24247147. doi:10.1176/appi.ps.201300256

Wang, S. V., Schneeweiss, S., Gagne, J. J., Evers, T., Gerlinger, C., Desai, R., & Najafzadeh, M. (2019). Using Real-World Data to Extrapolate Evidence From Randomized Controlled Trials. Clinical Pharmacology & Therapeutics, 105(5), 1156–1163. Retrieved from <Go to ISI>://WOS:000466750900020. doi:10.1002/cpt.1210

WHO. (2009). Guidelines for the Psychosocially Assisted Pharmacological Treatment of Opioid Dependence: Drug interactions involving methadone and buprenorphine. Retrieved from https://www.ncbi.nlm.nih.gov/books/NBK143177/

WHO. (2019). Guidelines for ATC classification and DDD assignment 2019. Retrieved from https://www.whocc.no/filearchive/publications/2019_guidelines_web.pdf

